# Microhomology-Mediated End Joining as a Novel Mechanism Underlying Androgen Insensitivity Syndrome

**DOI:** 10.1101/2025.06.24.25330003

**Authors:** Juliana Moreira Marques, Raquel Martinez Ramos, Nathália Da Roz D’Alessandre, Gabriela Der Agopian Guardia, Ana Caroline de Freitas Afonso, Barbara Leitao Braga, Mariana Ferreira de Assis Funari, Mirian Yumie Nishi, Paula Fontes Asprino, Sorahia Domenice, Pedro Alexandre Favoretto Galante, Berenice Bilharinho Mendonca, Rafael Loch Batista

## Abstract

**Significance Statement:** This study identifies microhomology-mediated end joining (MMEJ) as a novel mutational mechanism underlying a pathogenic deletion in the androgen receptor gene in a patient with complete androgen insensitivity syndrome. By mapping the genomic breakpoint at the nucleotide level, we demonstrate the presence of canonical features of MMEJ, thereby expanding the known mechanisms of genomic structural variation in *AR*. These findings underscore the importance of incorporating copy number variation (CNV) detection and breakpoint analysis into diagnostic workflows for 46,XY differences of sex development (DSD), enabling more accurate molecular classification and improved patient management.

**Background:** Copy number variations in the androgen receptor gene are an underrecognized cause of androgen insensitivity syndrome. Understanding their mutational mechanisms can improve AIS diagnosis and genotype–phenotype correlation.

**Objective:** To investigate microhomology-mediated end joining (MMEJ) as a mutational mechanism underlying a structural variant in the AR gene and to assess the contribution of AR CNVs to AIS through comparative gene burden analysis.

**Methods:** Whole-exome sequencing, Multiplex Ligation-dependent Probe Amplification, PCR, and Sanger sequencing were used to identify and refine a hemizygous deletion affecting exons 6–8 of the *AR* gene in a 46,XY individual with CAIS. Breakpoint mapping and local alignment were performed using R packages *Biostrings* and *GenomicRanges*. A literature and database review identified *AR* CNVs in AIS cases, which were compared to CNVs in the general population to assess AR-specific CNV burden.

**Results:** An accurate genomic analysis revealed an 8-bp microhomology region flanking the genomic breakpoint of the CNV event found in this CAIS patient, consistent with microhomology-mediated end joining event. Among 991 AIS cases, 49 harbored *AR* CNVs, significantly enriched compared to controls (OR = 4.59, P = 9.2 × 10⁻¹⁷). Exon 2 was the most frequently affected region and most strongly associated with CAIS.

**Conclusion:** This study provides the first molecular evidence that MMEJ can mediate pathogenic deletions in the *AR* gene. The significant enrichment of CNVs in AIS and their non-random distribution across functional domains support their role in disease pathogenesis and highlight the value of CNV-level analysis in the diagnostic evaluation of AIS.

## Introduction

The androgen receptor (AR) gene is essential for androgen signaling, encoding a nuclear receptor that regulates the physiological response to testosterone and dihydrotestosterone (DHT) (1). Located on chromosome Xq11-12, the (canonical form of) *AR* gene comprises eight exons, seven introns, and encodes a 920-amino-acid protein (2). Structurally, the AR protein shares characteristics with other nuclear receptors, containing three distinct functional domains: the N-terminal domain (NTD) (residues 1–555), the DNA-binding domain (DBD) (residues 556–623), the hinge domain (residues 624–665), and the ligand-binding domain (LBD) (residues 666–920) (2).

Pathogenic variants in the AR gene are the molecular basis of Androgen Insensitivity Syndrome (AIS), a condition characterized by resistance to androgen signaling (3). Clinically, AIS is classified into three phenotypic categories: complete AIS (CAIS), partial AIS (PAIS), and mild AIS (MAIS) (4). CAIS typically presents as an inguinal hernia or primary amenorrhea due to a complete inability to respond to androgens (5). PAIS manifests as undervirilized external genitalia, while MAIS is often associated with infertility or persistent gynecomastia in males (6, 7).

Structural variations, particularly copy number variations (CNVs), play a significant role in AIS but are often underappreciated in clinical practice. CNVs are genomic alterations that involve duplications or deletions of DNA segments, usually ranging from 1 kilobase to several megabases (8). These alterations can encompass entire genes or regulatory regions, affecting gene expression, dosage, and function. CNVs have been implicated in numerous genetic disorders, but their specific contribution to AIS remains incompletely understood (9).

The analysis of CNVs in the AR gene is particularly important, as the X-linked nature of the gene means that a single-copy deletion leads to complete gene loss in 46,XY individuals, resulting in severe androgen resistance. Large deletions within the AR gene, particularly those affecting the ligand-binding domain (LBD), have been associated with CAIS, whereas smaller deletions may result in PAIS or MAIS, depending on the extent of residual receptor function (10, 11). Identifying CNVs and their causal mechanisms in AR can provide crucial insights into genotype-phenotype correlations, improving diagnostic accuracy, prognostic predictions, and personalized treatment strategies. Among the mechanisms that give rise to CNVs, microhomology-mediated end joining (MMEJ) has emerged as a key DNA repair pathway responsible for non-recurrent genomic deletions (12). MMEJ is an error-prone form of double-strand break repair that relies on short (5–25 bp) regions of sequence (identity) homology to align and ligate DNA ends, typically resulting in deletions with microhomology flanking their junctions (13). Unlike homologous recombination, MMEJ does not require extensive sequence similarity. It has been implicated in structural variants underlying various genetic disorders and is particularly relevant to X-linked genes, where single-copy deletions have immediate phenotypic consequences in 46, XY individuals. Characterizing the sequence context at AR deletion breakpoints may provide insight into the mutational forces driving AIS and support the role of MMEJ in mediating clinically relevant structural variations.

In this study, we performed a molecular characterization of a large AR gene deletion identified in a patient with CAIS. Additionally, a comparative gene burden analysis was conducted to assess the prevalence of CNVs involving AR among affected individuals compared to the general population. We analyzed data from the AR mutation database, which included 49 CNVs from 991 individuals with AIS, and compared it to the gnomAD structural variant database (version SV v4.1.0), which contained 346 CNVs in 31,523 individuals serving as controls. The odds ratio (OR) was calculated to determine whether CNVs in AR are significantly enriched in AIS. This analysis aimed to assess whether CNVs in the *AR* region occur more frequently in AIS than in the general population, thus providing further evidence of pathogenic relevance of CNVs in AIS.

## Methods

### Whole Exome Sequencing (WES)

Whole-exome sequencing was performed using the SureSelect XT Human All Exon V6 Kit (Agilent). Libraries were sequenced on the Illumina HiSeq 2500 platform, generating paired-end reads (2 × 300 bp). Reads were aligned to the hg19 human genome assembly using BWA-MEM (RRID: SCR_010910). The aligned reads were sorted, and duplicate reads were flagged using the bamsort and bammark duplicates tools from biobambam2 (RRID: SCR_003308). Variant calling for single-nucleotide variants (SNVs) and small insertions/deletions (indels) was performed using FreeBayes (RRID: SCR_010761), and annotation was carried out with ANNOVAR (RRID: SCR_012821) (Wang, Li, and Hakonarson, 2010).

### Androgen Receptor (AR) Gene Sequencing

Genomic DNA was extracted from peripheral blood leukocytes using the proteinase K-SDS salting-out method. The entire AR gene (ENST00000374690) coding region, including exon-intron boundaries of exons 1 to 8, was amplified using previously published PCR primers (14,15). PCR products were sequenced using the ABI Prism BigDye Terminator Cycle Sequencing Ready Reaction Kit (Life Technologies, Carlsbad, CA) and analyzed on the ABI Prism Genetic Analyzer 3130XL (Life Technologies, Carlsbad, CA).

### Multiplex Ligation-dependent Probe Amplification (MLPA)

Structural variations in the AR gene, initially identified by PCR, were confirmed using the SALSA MLPA Reagent Kit (MRC-Holland) with the SALSA MLPA Probemix P074 AR, following the manufacturer’s protocol.

### Breakpoint and recombination analysis

Breakpoint analysis was performed to investigate the recombination mechanism underlying the AR gene deletion. Patient and reference AR genomic sequences (GRCh38/hg38) were compared using the UCSC Genome Browser and further analyzed in R (version 4.2.2) with Bioconductor packages Biostrings (version 2.66.0) and GenomicRanges (version 1.50.0). Additional dependencies included IRanges (version 2.32.0), XVector (version 0.36.0), and rtracklayer (version 1.56.1). Analyses were conducted under Bioconductor release 3.16.. A 200 bp region flanking each side of the predicted deletion breakpoint was extracted for local alignment. The pairwiseAlignment function from Biostrings was used to align the upstream and downstream sequences, enabling the identification of homologous and non-homologous segments involved in the recombination event. The exact breakpoint coordinates were mapped using GenomicRanges, allowing accurate annotation of the recombination site. Detection of short homologous sequences near the breakpoint supported inference of the DNA repair mechanism, such as microhomology-mediated end joining (MMEJ).

### Literature Review

A comprehensive literature review was conducted in PubMed from inception to August 10, 2024, without language restrictions. The search included the following keywords: (“Androgen Insensitivity Syndrome” OR “AIS” OR “46,XY Disorder of Sex Development” OR “46,XY DSD” OR “Testicular Feminization Syndrome” OR “XY Sex Reversal” OR “Male Pseudohermaphroditism”) AND (”Androgen Receptor” OR “AR gene” OR “NR3C4” OR “Dihydrotestosterone Receptor” OR “Testosterone Receptor”) AND (”Copy Number Variation” OR “CNV” OR “Gene Dosage” OR “Genomic Deletion” OR “Genomic Duplication” OR “Gene Copy Number” OR “Structural Variation” OR “Chromosomal Aberration” OR “Genomic Rearrangement” OR “Large Deletion” OR “Large Insertion” OR “Large Indel” OR “Gross Deletion”).

### Database Review

To investigate previously reported CNVs in the AR gene, genetic databases were reviewed, including the Androgen Receptor Database (http://androgendb.mcgill.ca/), the Human Gene Mutation Database (HGMD) (http://www.hgmd.cf.ac.uk/), and ClinVar (https://www.ncbi.nlm.nih.gov/clinvar/). Additionally, a secondary search was performed in the Genome Aggregation Database (gnomAD) (https://gnomad.broadinstitute.org) to ensure comprehensive data coverage. To assess the pathogenicity of CNVs in AR, a set of bioinformatic prediction tools was employed, including PolyPhen2, SIFT, REVEL, MetaLR, MetaRNN, MetaSVM, Mutation Assessor, MutationTaster, FATHMM, FATHMM-MKL, LRT, M-CAP, and DAAN (https://franklin.genoox.com/clinical-db/home).

### Gene Enrichment Analysis

To assess whether copy number variations (CNVs) in the *AR* gene are significantly enriched in individuals with Androgen Insensitivity Syndrome (AIS), we performed a comparative gene burden analysis using two independent datasets. The AIS cohort comprised 991 individuals, among whom 49 carried CNVs in *AR*, as identified through curated mutation databases. For comparison, structural variant data from 31,523 individuals in the gnomAD database (version SV v4.1.0) revealed 346 CNVs in *AR*.

An odds ratio (OR) was calculated to estimate the relative likelihood of *AR* CNVs occurring in AIS cases versus the general population. Fisher’s exact test was used to evaluate statistical significance, with a two-tailed P value *p*< 0.05 considered significant.

Genomic coordinates of the CNVs were mapped to the human reference genome and visualized using Python. A heatmap was generated to display the frequency of CNVs affecting each exon/intron combination. In parallel, CNVs were plotted across the genomic coordinates of the *AR* locus with color coding based on phenotype, allowing positional correlation with functional domains of the gene and potential phenotype-specific clustering. This analysis aimed to quantify the burden of *AR* structural variation in AIS and to determine whether CNVs in this gene represent a non-random, disease-associated event.

### Ethics Statement

Written informed consent was obtained from the patient for genetic and laboratory studies. This study was approved by the local ethical committee (CAAE: 66367717.6.0000.0068).

### Data Availability

The datasets analyzed during the current study are publicly available on HGMD (https://www.hgmd.cf.ac.uk/), the androgen receptor mutations database (http://androgendb.mcgill.ca/), and the Genome Aggregation Database (gnomAD, https://gnomad.broadinstitute.org).

## Results

### Clinical and Hormonal Findings

An individual in their early 20s, assigned female at birth, with a history of primary amenorrhea, presented for evaluation after multiple unsuccessful attempts to induce menstruation with progestin therapy during adolescence. They also had a history of a right-sided inguinal hernia in early childhood, which had been managed conservatively. On physical examination, she was in good general condition (BMI 25 kg/m²), with typical female external genitalia, Tanner stage M5 breast development, and markedly sparse pubic hair. No gonads were palpable in the inguinal region.

Laboratory testing revealed elevated total testosterone, increased luteinizing hormone (LH), and normal follicle-stimulating hormone (FSH) levels (Table 1). Cytogenetic analysis confirmed a 46,XY karyotype. Pelvic magnetic resonance imaging demonstrated absence of the uterus, cervix, and upper vagina, along with bilateral gonadal-like structures in the iliac fossae.

**Table 1.**
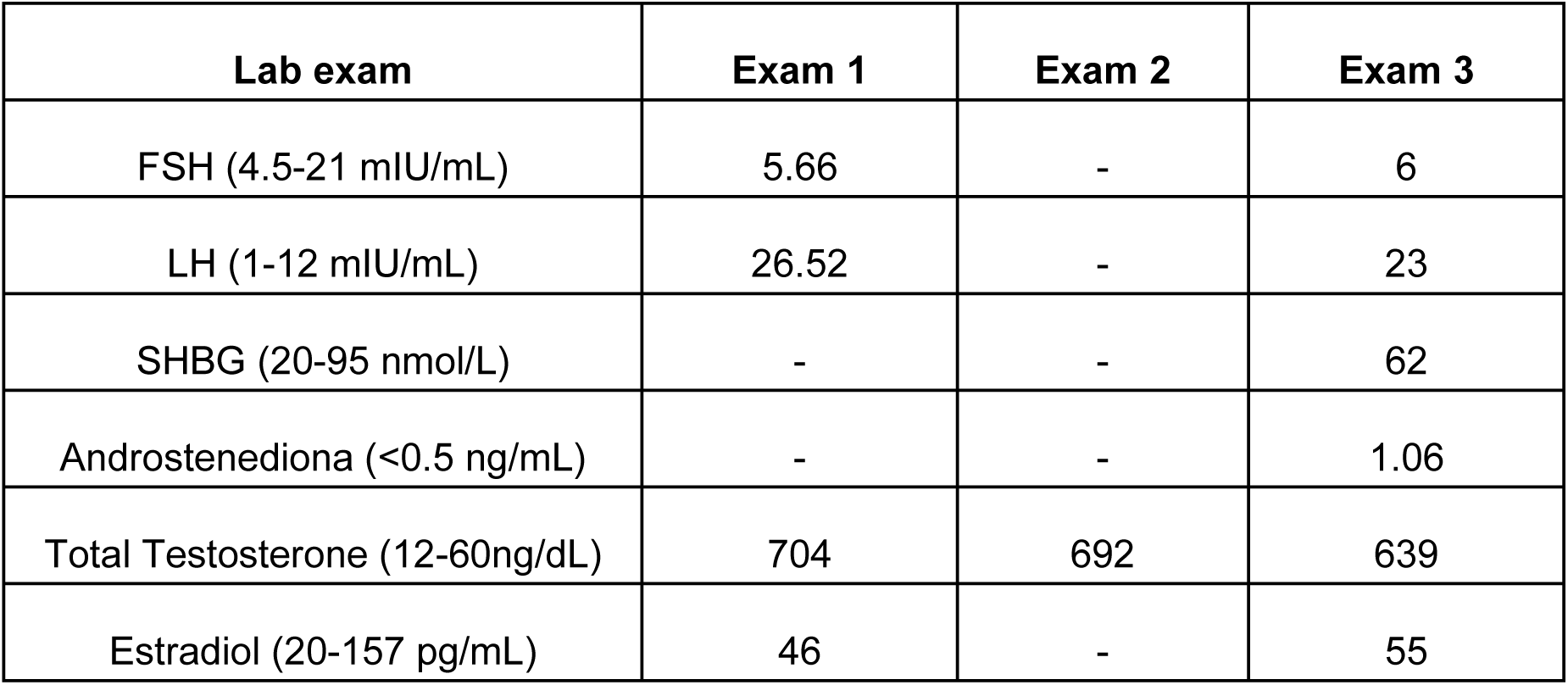
Hormonal profile of the patient during diagnostic evaluation. Abbreviations: FSH – *Follicle-Stimulating Hormone*; LH – *Luteinizing Hormone*; SHBG – *Sex Hormone-Binding Globulin*.

### Molecular Characterization of a Deletion in the AR Gene

Whole Exome Sequencing (WES) identified a deletion of approximately 2,008 base pairs on chromosome X (chrX:66,941,674–66,943,682; GRCh38), encompassing exons 6, 7, and 8 of the *AR* gene (Figure 1A). These exons encode the majority of the ligand-binding domain (LBD) of the androgen receptor. No additional pathogenic variants were detected in other genes associated with disorders of sex development (DSD).

**Figure 1.**
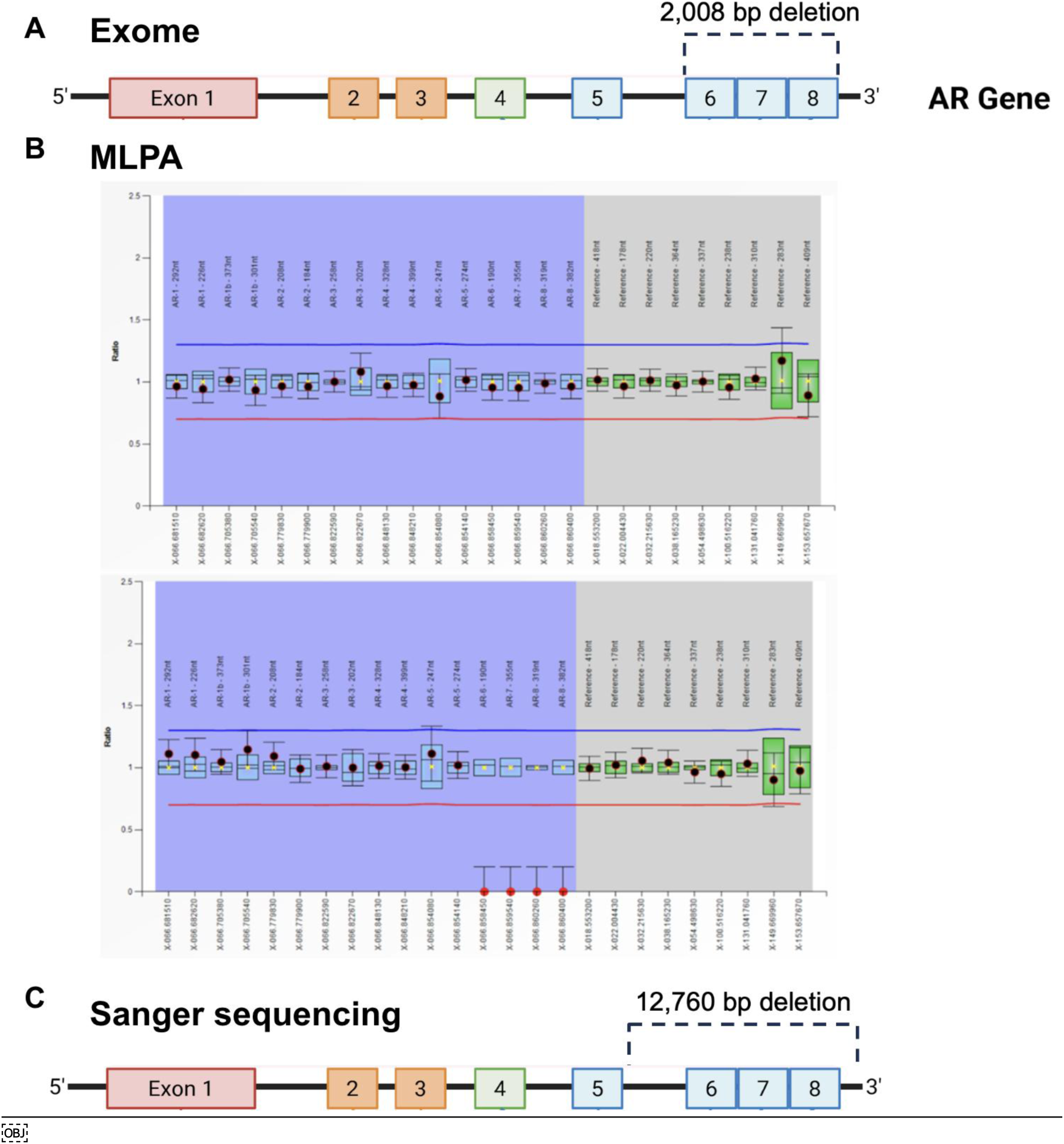
Identification and refinement of deletions in the AR gene using exome sequencing, MLPA, and Sanger sequencing. **(A)** Schematic representation of the AR gene exons (1–8), with a 2,008 bp deletion initially identified by exome sequencing, spanning exons 6 to 8. **(B)** MLPA (Multiplex Ligation-dependent Probe Amplification) analysis identified a deletion encompassing exons 6–8 of the AR gene. Probe signal intensities from the patient were compared to reference samples to assess copy number variation. The blue and red horizontal lines represent arbitrary thresholds set at approximately ±0.3 from the average probe value across reference samples. Blue and green boxes correspond to the 95% confidence intervals (CI) for each probe in the reference group. Probes in the blue-shaded zone target exonic regions of interest. Probes in the gray-shaded zone serve as internal controls. Black dots represent patient probe ratios within the 95% CI; red dots fall outside this interval, suggesting altered copy number. **(C)** Refinement of the deletion breakpoints by Sanger sequencing revealed a larger deletion of 12,760 bp encompassing the same exons, confirming and extending the initial finding from exome data.

To validate and refine the identified structural variant, we employed Multiplex Ligation-dependent Probe Amplification (MLPA), which revealed reduced probe signal intensity for exons 6–8, consistent with a deletion in this region (Figure 1B). PCR and Sanger sequencing further confirmed the deletion and defined its boundaries with higher resolution, revealing a 12,760 bp deletion that spanned the same exons (Figure 1C). This discrepancy in size compared to the WES-based call reflects the limited breakpoint resolution of short-read exome data. The combined approach confirmed the loss of the *AR* ligand-binding domain due to a large genomic deletion.

To investigate the mechanism underlying the AR gene deletion, breakpoint analysis was performed using local sequence alignment of upstream and downstream flanking regions. The deleted segment was bordered by two 8 bp microhomologous sequences located 3 bp upstream of the 5′ breakpoint and 32 bp downstream (in reverse complement) of the 3′ breakpoint, suggesting the use of microhomology-mediated end joining (MMEJ) as the most likely DNA repair mechanism (Figure 2A). This was further supported by small insertions and retained nucleotides flanking the breakpoint junction — a hallmark of MMEJ-mediated repair.

**Figure 2.**
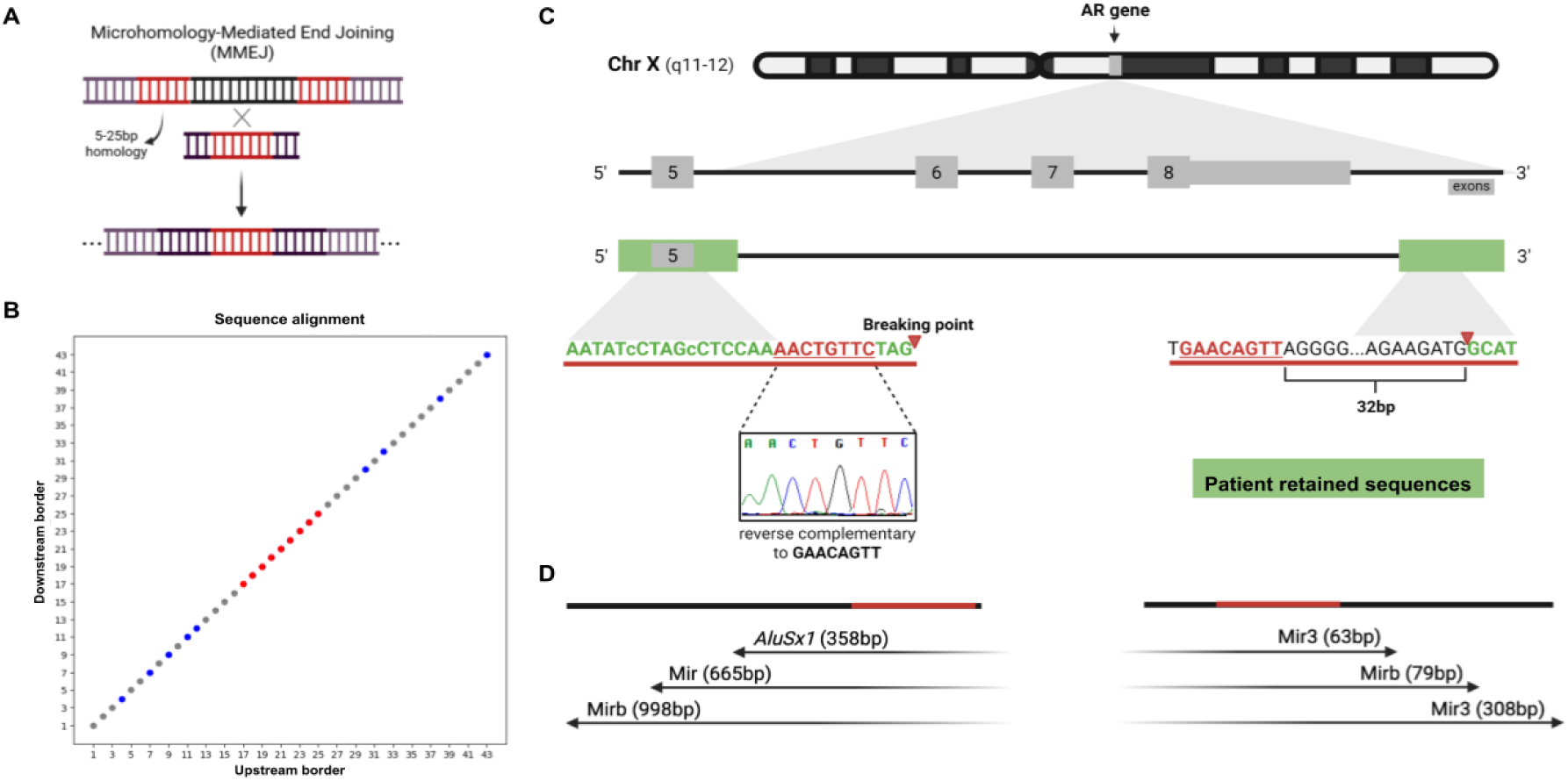
Microhomology-mediated end joining (MMEJ) as the mechanism underlying an AR gene deletion in a patient with androgen insensitivity syndrome (AIS). **(A)** Schematic of MMEJ, a DNA repair mechanism that utilizes short homologous sequences (5– 25 bp) to mediate end joining, often resulting in deletions. **(B)** Dot plot alignment of upstream and downstream breakpoint borders reveals a region of microhomology (red), consistent with MMEJ-mediated repair. **(C)** Genomic context of the AR gene on chromosome X, showing a deletion spanning exons 6 to 8. A microhomology of 8 bp (GAACAGTT) was identified 3 bp upstream of the 5′ breakpoint and 32 bp downstream (within the deleted region) on the 3′ side in reverse complement orientation, consistent with a microhomology-mediated end joining (MMEJ) mechanism. (D) *Alu* and MIR elements are present near the breakpoints but distant from the junction, suggesting an indirect role in promoting genomic instability.

Local pairwise alignment confirmed the microhomology region and the absence of extensive homology required for homologous recombination (Figure 2B). Additionally, short insertions retained in the junction site (Figure 2C, lower case nucleotides), together with the offset position of the microhomology relative to the breakpoints, are consistent with template alignment, microhomology recognition, and resection-dependent repair, as described in MMEJ. Repetitive elements from the *Alu* and MIR subfamilies—both members of the SINE (Short Interspersed Nuclear Elements) family— were identified near both breakpoints (Figure 2D), although they were located at considerable distances from the deletion boundaries. While no direct involvement was observed, the presence of these elements may have contributed to local genomic instability or structural susceptibility to breakage and repair via MMEJ.

### Enrichment of CNVs in AIS Compared to Controls

To assess the pathogenic relevance of AR CNVs, we compared the frequency of structural variants in 991 individuals with AIS (49 CNVs) to that in 31,523 individuals from the gnomAD database (346 CNVs). The analysis revealed a significant enrichment of CNVs in AIS cases (OR = 4.59, *P* = 9.2 × 10⁻¹⁷), reinforcing the contribution of large deletions to the molecular etiology of AIS. A total of 49 CNVs affecting the *AR* gene were compiled and mapped by genomic region and clinical phenotype (Figure 3A, 3B, Table 2). The most frequently affected region was exon 2, with 12 independent cases, followed by exons 1 and 1–8 deletions. Heatmap analysis revealed a non-random distribution of CNVs, with clustering in exons encoding the DNA- and ligand-binding domains (Figure 3A). When stratified by phenotype, 84% of the CNVs were associated with CAIS, 14% with PAIS, and 2% with MAIS. Notably, most deletions associated with CAIS involved exon 2 or the entire 3′ portion of the gene, whereas PAIS-linked CNVs were more frequently located in intron 2 or partially spanned exons 5–7, suggesting a possible genotype–phenotype correlation influenced by CNV extent and location.

**Figure 3.**
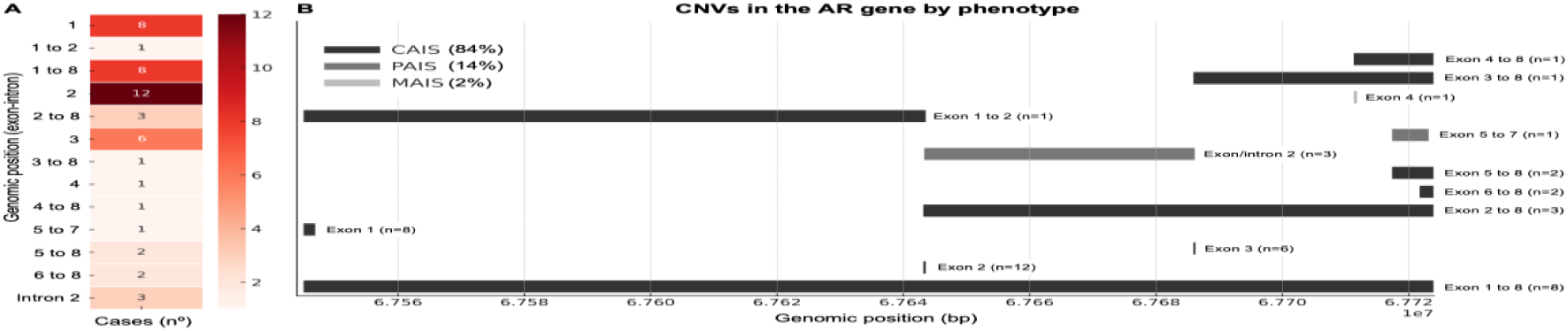
Genomic distribution and phenotypic correlation of CNVs in the AR gene. **(A)** Heatmap showing the number of reported CNV cases affecting specific regions of the AR gene. The highest concentration of CNVs was observed at exon 2 (n=12), followed by exons 1 (n=8) and 1–8 deletions (n=8), supporting a recurrent breakpoint hotspot. **(B)** Genomic locations of CNVs mapped along the AR gene, color-coded by associated phenotype: CAIS (dark gray), PAIS (medium gray), and MAIS (light gray). Exon 2 deletions represent the most frequent event (n = 12), predominantly associated with the most severe phenotype, complete androgen insensitivity syndrome (CAIS). The relative proportions of CAIS (84%), PAIS (14%), and MAIS (2%) are indicated.

**Table 2.** Copy Number Variations (CNVs) in the AR Gene in Individuals with Androgen Insensitivity Syndrome (AIS).

| Phenotype | Number of Cases | Variant Type | Affected Region | Reference |
| --- | --- | --- | --- | --- |
| CAIS | 1 | deletion | exon 1-8 | (27) |
| CAIS | 1 | deletion | exon 1-8 | (28) |
| CAIS | 1 | deletion | exon 1-8 | (22) |
| CAIS | 2 | deletion | exon 1-8 | (29) |
| CAIS | 2 | deletion | exon 2 (7028 bp) | (30) |
| CAIS | 3 | deletion | exon 1-8 | (31) |
| CAIS | 2 | deletion | exon 2-8 | (32) |
| CAIS | 2 | deletion | exon 2 | (33) |
| CAIS | 2 | deletion | exon 3 | (31) |
| PAIS | 1 | deletion | exon 3 | (22) |
| CAIS | 1 | deletion | exon 4-8 | (34) |
| CAIS | 1 | deletion | exon 3-8 | (34) |
| MAIS | 1 | deletion | exon 4 | (21) |
| CAIS | 1 | deletion | exon 1-2 | (35) |
| CAIS | 2 | deletion | exon 5-8 | (36) |
| CAIS | 1 | deletion | exon 6-8 | (44) |
| PAIS | 1 | deletion | intron 2 (>6kb) | (36) |
| CAIS | 1 | deletion | exon 3 | (37) |
| CAIS | 2 | deletion | exon 3 | (38) |
| PAIS | 2 | deletion | intron 2 (6 kb) | (39) |
| CAIS | 1 | duplication | exon 2 | (38) |
| CAIS | 1 | deletion | exon 2 | (33) |
| PAIS | 1 | deletion | exon 5-7 | (19) |
| CAIS | 2 | deletion | exon 2 | (40) |
| CAIS | 2 | deletion | exon 2 | (40) |
| CAIS | 7 | indel | exon 1 del287ins77 | (41) |
| CAIS | 1 | deletion | exon 2-8 | (18) |
| CAIS | 1 | duplication | exon 2 | (17) |
| PAIS | 1 | deletion | exon 1 | (18) |
| PAIS | 1 | deletion | exon 2 | (18) |
| CAIS | 1 | deletion | exon 6-8 |  |

### Genomic mapping of CNVs and AIS phenotype association

To further explore genotype–phenotype correlations, we compiled reported CNVs in the *AR* gene from the literature and databases and mapped their distribution across the gene structure. As shown in Figure 3B, deletions affecting exon 2 were the most frequently reported (n = 12), followed by deletions encompassing the entire gene or large segments (e.g., exons 1–8 or 2–8, each with n = 8). These regions represent recurrent hotspots for structural variation. Notably, most CNVs clustered in the 5′ half of the gene, with exon 2 and intron 2 emerging as high-density zones for genomic rearrangements.

When analyzed by associated phenotype (Figure 3A), the majority of CNVs were observed in individuals with complete AIS (CAIS, 84%), while fewer were associated with partial AIS (PAIS, 14%) or mild AIS (MAIS, 2%). Deletions encompassing multiple exons—especially exons 2 to 8 or exons 1 to 8—were almost exclusively linked to CAIS. In contrast, smaller or intronic deletions, such as those involving intron 2 or exons 5–7, were more frequently found in PAIS or MAIS, suggesting partial retention of AR function. These findings reinforce the genotype–phenotype correlation between CNV extent and clinical severity and support the diagnostic value of mapping CNVs at exon-level resolution.

## Discussion

Identifying the molecular etiology of 46,XY DSD is essential for personalized care (14). In CAIS, early diagnosis facilitates timely decisions regarding gonadal management and gender-affirming care (15). In partial AIS (PAIS), the underlying variant may influence surgical planning, fertility counseling, and long-term follow-up (6, 16). Integrating molecular and clinical data is crucial to improving outcomes in AIS.

This study contributes to the growing recognition that CNVs in the androgen receptor gene (AR) are a significant, and often underrecognized, cause of AIS. We describe a patient with a 46,XY karyotype, classical CAIS phenotype, and a large deletion encompassing exons 6–8 of *AR*, corresponding to the ligand-binding domain (LBD). The diagnosis was confirmed by orthogonal methodologies, WES, MLPA, PCR, and Sanger sequencing, highlighting the need for CNV-sensitive methods when routine sequencing fails to detect pathogenic variants.

Recently, a novel duplication involving exon 2 of the *AR* was reported in a patient with CAIS, which was missed by conventional sequencing but successfully detected through a WES pipeline with integrated CNV Calling (17). Their findings underscore the need for routine implementation of CNV-sensitive approaches, including WES with dedicated CNV-calling algorithms and MLPA, especially in patients with strong clinical suspicion but no identifiable single-nucleotide variants. Our case aligns with this perspective and further demonstrates that not only deletions, but also duplications, should be considered in the diagnostic workflow for AIS.

A review of the literature identified 49 previously reported CNVs in the AR gene, most of which were associated with CAIS. These typically involved large deletions spanning multiple exons. Although less frequent, CNVs have also been reported in individuals with PAIS or even MAIS (18–22), indicating that structural variants may underlie a broader spectrum of AIS phenotypes. Supporting the clinical relevance of these findings, a gene burden analysis demonstrated a significant enrichment of AR CNVs in AIS patients compared to the general population (OR = 4.59; P = 9.2 × 10⁻¹⁷). Despite advances in sequencing technologies, structural variants remain underdiagnosed in AIS. Our findings emphasize the importance of integrating CNV analysis—through MLPA, WES-based algorithms, or genome-wide arrays— into routine diagnostic workflows, particularly in unsolved cases with high clinical suspicion.

Importantly, this is the first report to implicate MMEJ as the mechanism underlying a pathogenic AR deletion. Genomic deletions result from different DNA repair pathways activated in response to double-strand breaks (DSBs) (23). While homologous recombination (HR) uses long identical DNA templates for accurate repair, non-homologous end joining (NHEJ) and MMEJ operate without a template and are more error-prone (24). MMEJ relies on short microhomologous sequences (5–25 bp) to align DNA ends before ligation, frequently resulting in the loss of intervening DNA (13). It has been increasingly recognized as a source of non-recurrent deletions, particularly in X-linked genes, where single-copy loss is sufficient to disrupt function in 46,XY individuals.

In this CAIS patient, a genomic sequence alignment involving the flanking region of the deletion event revealed an 8 bp of microhomology, with no evidence of long repeats or homologous sequences, supporting MMEJ as the likely repair mechanism. This aligns with canonical features of MMEJ: CtIP-mediated resection, alignment via short homology, Polθ-mediated synthesis, and LIG3/LIG1 ligation. Our findings provide the first direct breakpoint-level evidence of MMEJ in AR and offer a molecular framework for interpreting future deletions based on their repair signatures.

Although MMEJ-mediated deletions have not been previously reported in the *AR* gene, mobile element insertions—particularly LINE-1—have been reported in the AR 5’UTR region in individuals with PAIS (25, 26). These insertions exhibit classical hallmarks of retrotransposition, such as target site duplications and poly(A) tails, and were associated with significant impairment of AR transcription. While mechanistically distinct from MMEJ, these findings reinforce that non-canonical mutational mechanisms may contribute to monogenic disorders and highlight the vulnerability of the AR gene to structural rearrangements.

In conclusion, this study expands the mutational spectrum harboring the *AR* gene by identifying MMEJ as a novel mechanism underlying pathogenic deletions in AIS. Precise breakpoint event mapping offers mechanistic insight and enhances genomic variant classification. These findings support the incorporation of genomic structural variant detection and repair-pathway analysis into clinical diagnostics, advancing precision medicine in 46,XY DSD.

